# Macular Oxygen Saturation in Glaucoma Using Retinal Oximetry of Visible Light Optical Coherence Tomography

**DOI:** 10.1101/2023.12.20.23300300

**Authors:** Jingyu Wang, Natalie Sadlak, Marissa G. Fiorello, Manishi Desai, Ji Yi

**Affiliations:** Department of Ophthalmology, School of Medicine, Johns Hopkins University, Baltimore, MD, USA; Department of Ophthalmology, Boston Medical Center, Boston, MA, USA; Department of Medicine, Boston University School of Medicine, Boston Medical Center, Boston, MA, USA; Department of Biomedical Engineering, Johns Hopkins University, Baltimore, MD, USA

**Author notes:** Corresponding author: Ji Yi.

**Keywords:** visible light optical coherence tomography, retina oximetry, glaucoma

## Abstract

**Purpose:** Oxygen saturation (sO_2_) plays a critical role in retinal pathophysiology, especially at the macula, which undergoes significant energy consumption. While macular damage has been suggested to be involved in early-stage glaucoma, there has been no report to date on non-invasive macular sO_2_ in glaucoma. Therefore, we conducted this study to compare macular sO_2_ associated with other clinical measurements between normal and glaucoma subjects and evaluate whether there are significant differences.

**Method:** This is a cross-sectional study. We used visible light optical coherence tomography (VIS-OCT) for retinal oximetry in perifoveal vessels. The subjects from groups of normal, suspect/pre-perimetric glaucoma (GS/PPG) and perimetric glaucoma (PG) were scanned using VIS-OCT in the macular region with a sampling density of 512×256 in an area of 5×5 mm^2^. 48 eyes (16 normal, 17 GS/PPG and 15 PG) were included for the analysis. For each eye, we measured the sO_2_ of arterioles (AsO_2_), venules (VsO_2_), and calculated the difference between arterioles and venules (A-V sO_2_=AsO_2_-VsO_2_), oxygen extraction (OE=(AsO_2_-VsO_2_)/AsO_2_ ×100%). Additionally, we included Zeiss Cirrus OCT scans and 24-2 visual field test (VFT) for clinical benchmark. One-way ANOVA was used to compare the differences among the three groups. Spearman correlation tests were used for correlation sO_2_ markers to standard metrics including the thickness of ganglion cell layer and inner plexiform layer (GCL+IPL), circumpapillary retinal nerve fiber layer (cpRNFL) and mean deviation (MD) in VFT.

**Result:** Significant differences were found among three groups for all VIS-OCT, Zeiss OCT, and VFT variables. Macular AsO_2_, A-V sO2, OE decreased, and VsO_2_ increased along with severity. Macular AsO_2_ and A-V sO_2_ were statistically correlated with GCL+IPL and cpRNFL in all eyes, as well as only PG eyes. Within PG eyes, the correlation between AsO_2_ and GCL+IPL is dominant in more damaged lower hemifield.

**Conclusion:** The GS/PPG and PG subjects had significantly higher macular VsO_2_, lower A-V sO_2_ and OE indicating less oxygen consumption. The sO_2_ measured by retinal oximetry of VIS-OCT can be a potential metric for the early diagnosis of glaucoma.

## Introduction

Glaucoma is an optical neuropathy, causing irreversible blindness and impacting millions worldwide^1,2^. The clinical hallmark of glaucoma is the loss of retinal ganglion cells (RGCs) and nerve fiber layer (RNFL), leading to chronic vision deterioration. Vision loss in glaucoma is permanent and irreversible, emphasizing the importance of early detection for prevention and clinical management^3–6^.

While the initial insult occurs at optic nerve head, it retrogradely damages the axon and soma of RGCs^7^. Because the macula contains >30% of total RGCs in retina^8–10^, changes associated with RGC damage may be detected in macular region. Indeed, evidence supported that early stages of glaucoma may involve macular RGC loss that warrants close attention^8,11^. Thinning of macular ganglion cell complex is associated with glaucoma^12^. Optical coherence tomography angiography found reduced macular superficial capillary density in glaucoma eyes, and the rate of capillary density can indicate glaucoma worsening in some cases^13–15^. Using a 10-2 VFT with denser 2° sampling density, a recent study showed central visual damage in over 30% of ocular hypertensive and glaucomatous eyes while the conventional 24-2 VFT appeared normal^16^.

The retina is one of the highest oxygen-consuming tissues, with RGCs being the major consumer in inner retina, perfused by retinal circulation^17,18^. Therefore, we hypothesize the macular inner retina oxygen extraction can assess RGC function, allowing the measurement of reduced oxygen extraction in the macular region to indicate the damage or loss of RGCs. By measuring blood sO_2_ from parafoveal arterioles and venules extended from superior and inferior arcades towards macula, we can assess macular oxygen consumption by arteriovenous sO_2_ (A-V sO_2_) and oxygen extraction.

To non-invasively measure macular vessel sO_2_, visible light optical coherence tomography (VIS-OCT) is used in this study. VIS-OCT is an emerging retinal imaging method that uses visible light instead of conventional near infrared (NIR), resulting in much higher axial resolution and significant absorption contrast between oxygenated and deoxygenated hemoglobin^19–21^. This makes retinal oximetry using VIS-OCT feasible for quantifying sO_2_ in human eyes^22–24^. Compared to the existing Oximap based on multi-wavelength fundus images^25–27^, VIS-OCT provides unattainable depth-resolved capabilities, allowing for more precise measurement of sO_2_ in macular vessels and even capillaries ^28^.

Here, the goal of this presented study is to utilize retinal oximetry of VIS-OCT to measure macular sO_2_ in normal, GS/PPG and PG groups, evaluate the feasibility of sO_2_ to be an early biomarker to differentiate among these three groups, and explore the association between sO_2_ and glaucoma-represented parameters of GCL+IPL and cpRNFL.

## Method

### Human subjects

The Institutional Review Board of Boston Medical Center reviewed and approved this study, ensuring compliance with the Health Insurance Portability and Accountability Act. The study took place from March 2019 to January 2020. All the subjects were provided with the tenets of Declaration of Helsinki, and written informed consent was obtained from each participant. We recruited the subjects in control group through the Boston Medical Center Optometry clinics, and the clinical subjects from in BMC eye clinics during their standard of care visit.

### Clinical examination

Prior to the examination, the pupils were dilated by Tropicamide. Subjects underwent tonometry, stereoscopic optic disc assessment and clinical OCT imaging (Cirrus, Zeiss, Jena, Germany). We recorded the quantitative results including cpRNFL and GCL+IPL, cup to disc ratio (CDR) and vertical cup-disc ratio (VCDR) from Cirrus OCT scans. For all the clinical subjects, central 24-2 threshold visual field tests (VFTs) were conducted with mean deviation (MD) and pattern standard deviation (PSD). We evaluated the cataracts using the Lens Opacification System II based on color and opalescence utilizing a 4-point grading system with an increasing number consistent with increasing maturity. After clinical and ophthalmic examinations, a trained technician imaged the subjects using the dual-channel VIS-OCT system with both eyes if eligible.

We acquired the images centered at the fovea using a custom-built dual-channel VIS-OCT system ^29^ with a raster scanning of 512 A-lines by 256 B-scans covering an area of 5×5 mm^2^. The illumination wavelengths were 545 to 580 nm for the visible channel, and 800 to 880 nm for the NIR channel. The illumination power on pupil was less than <0.25 mW (VIS) and 0.9 mW (NIR), respectively, meeting the safety standards of the ANSI for ophthalmic instrument. Details of the calculation can be seen in the supplementals of our previous publication^30^. We used a tunable lens to correct the spherical refractive error. We used the fellow eye for fixation with an external target. The start of the imaging used the NIR channel for alignment and focus tuning. After confirming desired locations with the best preview image quality, we initiated the dual-channel acquisition. The A-line rate of camera is 50 kHz with an exposure time of 19.1 us. The total acquisition time for one raster scan was 2.62s.

### Image processing and sO_2_ calculation

After acquisition, volumetric 3D data from both channels were produced by DC spectrum removal, k-space resampling, dispersion compensation and fast Fourier transform. Detailed processing steps are described in the past work^24^. In NIR dataset, we first segmented the layer of retinal pigment epithelium (RPE) by detecting the location of maximum intensity in each A-line, then detected the location of maximum gradient above the RPE layer for ILM boundary. We used outlier detection and 3^rd^ order polynomial curve fitting to smooth the segmentation of RPE and ILM, then registered them in VIS data. We generated the *en face* vessel map by averaging the space between RPE and 20 pixels above RPE for the maximum contrast, then manually chose the vessels as vessel ROI mask.

Using short time Fourier transform, 11 Gaussian windows swept the interferogram for four-dimensional (4D) dataset *I* (*x, y, z, λ*). A-lines within vessel ROIs were averaged for spectrum I (*z, λ*), which is further normalized by an averaged spectrum from non-vascular RNFL. With the known ILM boundary, we located the vessel bottom and averaged the spectrum in depth from 5 pixels above the vessel bottom and 10 pixels below to generate a single spectrum for each vessel ROI.

A least-square fitting on the extracted spectra calculates sO_2_ for each vessel ROI using the algorithm below as in **Figure 1**

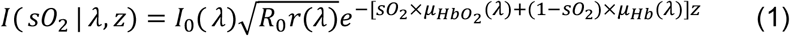

where *I*_0_( *λ*) is the spectrum of light source; *R*_0_ is an assumed constant for the reflectance of reference arm; *r*(*λ*) (dimensionless) is the reflectance at the vessel wall, modelled by a power law *r*(*λ*) = *Aλ*^−*α*^, with A being a dimensionless constant and modelling the decaying scattering spectrum from the vessel wall. The optical attenuation coefficient *μ* is determined by the coefficients of absorption *μ*_*a*_ and scattering *μ*_*s*_, where *μ*(*λ*) = *μ*_*a*_ (*λ*) + *Wμ*_*s*_(*λ*). *W* is the scaling factor for the scattering coefficient which was 0.2 used here. After obtaining the sO_2_ for all vessels, we classified them into the sO_2_ of arterioles (AsO_2_) and venules (VsO_2_) based on the alternation pattern and values. We defined sO_2_ difference between arterioles and venues (A-V sO_2_=AsO_2_-VsO_2_) and oxygen extraction (OE= (AsO_2_ – VsO_2_)/AsO_2_*100%) for further investigation.

**Figure 1.**
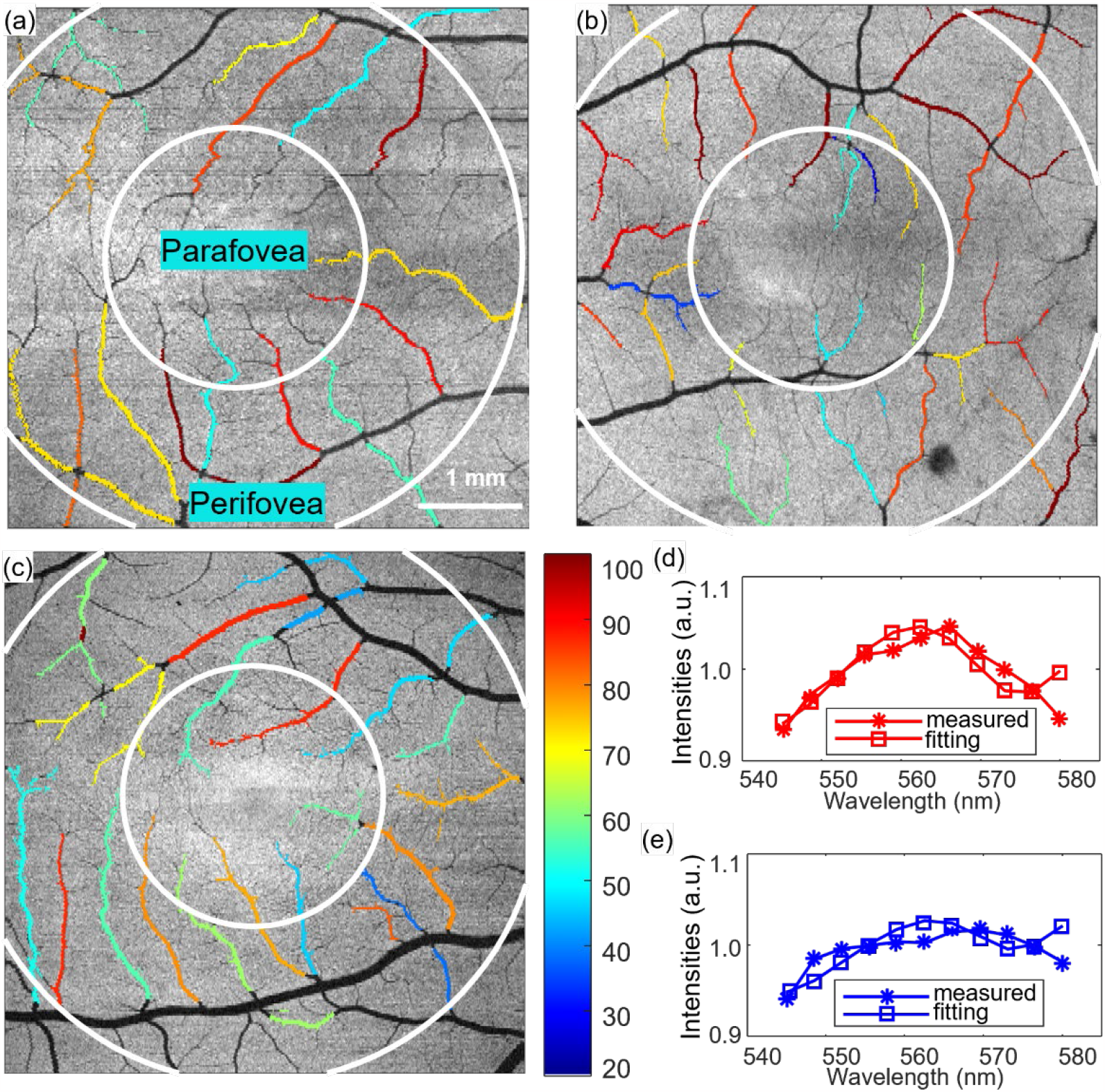
sO_2_ maps and sO_2_ calculation (a-c) sO_2_ map of a right, left and right eyes from the Normal, GS/PPG, and PG subjects. (d-e) Fitting for calculating sO_2_ of arterioles and venules.

### Study group definition

We defined a normal eye as one with a normal-appearing optical nerve head, assessed by stereoscopic optic disc photograph at the point of care (subjective assessment of vertical cup-disc ratio), and intraocular pressure (IOP) ≤ 22 mmHg. We defined the GS/PPG as an optical disk potentially presenting glaucomatous optical neuropathy after stereoscopic optic nerve examination. The visual field was normal by the 24-2 VFT threshold test, and the glaucoma hemifield test was within normal range or borderline. The index of pattern standard deviation was less than 5%. We defined the PG as an optic disc compatible with glaucoma. The subjects had an abnormal visual field, including the result of glaucoma hemifield outside the normal limit or the index of pattern standard deviation was less than 0.05%.

### Inclusion and exclusion criterion

We included subjects over 40 years old and with visual acuity better than 20/40 after correction. For the experimental group, we only included the patients diagnosed with primary open angle glaucoma (POAG) or potential POAG, excluding other ocular conditions such as primary angle closure glaucoma (PACG), intraocular surgeries (except for uncomplicated cataract surgery), history of diabetic retinopathy, vascular occlusion, macular degeneration, macular edema, hereditary retinal degeneration, uveitis, and other retinal conditions. We excluded the subjects from both groups due to fixation failure and low image quality that rendered them unattainable for segmentations. Subjects with severe cataracts graded more than 2+ were also excluded, considering the sensitivity of visible light to cataract and lens changes.

### Statistical analysis

We evaluated the categorical variables by Fisher-Freeman-Halton exact test and Chi-square test. For continuous variables, we applied one-way ANOVA across the three groups, followed by multiple comparisons. We employed the t-test for two group comparisons. We conducted the Receiver operating characteristic (ROC) analysis to assess detection accuracy based on the univariate logistic regression. We did the Spearman correlation test among the variables to get the correlation coefficients and p-values. It is noted as significant with bold when the *p* value was less than 0.05.

## Results

### Demographics

A total of 48 eyes of 35 subjects were included in the analysis:16 eyes of 10 normal subjects, 17 eyes of 12 GS/PPG subjects and 15 eyes of 13 GS subjects. The primary reasons for exclusion were server cataracts and fixation failure, which resulted in incomplete *en face* macular images and the inability to segment retinal layers for subsequent sO_2_ analysis. The demographic was presented in **Table 1**. No significant differences were observed among the three groups regarding eye side, age, gender, race, and ethnicity.

**Table 1.** Demographic of subjects.

|  | Normal | GS/PPG | PG | P-value |
| --- | --- | --- | --- | --- |
| Subjects(n) | 10 | 12 | 13 | - |
| Eyes (OD/OS) | 8/8 | 8/9 | 8/7 | 0.939 (a) |
| Age (mean± standard deviation) | 61.0±13.4 | 63.3±10.3 | 63.6±12.9 | 0.865 (b) |
| Gender (female/male) | 5/5 | 6/6 | 7/6 | 0.976(a) |
| Race (AA/White/N.A.) | 5/3/2 | 7/4/1 | 6/4/3 | 0.936 (c) |
| Ethnicity (non-Hispanic/Hispanic/N.A.) | 7/2/1 | 11/1/0 | 11/1/1 | 0.727 (c) |

AA: African American. N.A., not applicable. (a) Chi-square test. (2) One way ANOVA test. (c) Fisher-Freeman-Halton exact test

### Key measurements

Ocular measurements among three groups are summarized in **Table 2**. In visual field testing (VFT), the differences between GS/PPG and PG are significant for both 24-2 MD and 24-2 PSD. Additionally, all measurements obtained from Cirrus and VIS-OCT are all significantly different among three groups.

**Table 2.** Characteristics of ocular measurements from eyes among three groups.

|  | Normal<br>(n=16) | GS/PPG<br>(n=17) | PG<br>(n=15) | P-value |
| --- | --- | --- | --- | --- |
| Ophthalmic Exam |  |  |  |  |
| Cataract (Yes/No/N.A.) | 9/7/0 | 10/6/1 | 13/0/2 | **0.013** (a) |
| Pseudophakic lens (Yes/No/N.A.) | 3/13/0 | 4/12/1 | 0/13/2 | 0.181 (a) |
| IOP (mean ± Std mmHg) | 15.4 ± 2.3 | 14.8 ± 3.6 | 13.5 ± 3.8 | 0.321 (b) |
| Cylinder (mean ± Std) | 1.00 ± 0.71 | 0.46 ± 0.37 | 0.83 ± 0.53 | 0.051 (b) |
| Visual field test |  |  |  |  |
| 24-2 MD (mean ± Std dB) | - | -0.59 ± 0.97 | -7.60 ± 4.67 | **<0.001** (c) |
| 24-2 PSD (mean ± Std dB) | - | 1.91 ± 0.39 | 7.71 ± 4.63 | **<0.001** (c) |
| Zeiss OCT |  |  |  |  |
| GCL+IPL (mean ± Std μm) | 78.1 ± 8.9 | 73.8 ± 8.2 | 64.0 ± 11.3 | **<0.001** (b) |
| cpRNFL (mean ± Std μm) | 90.9 ± 10.0 | 85.4 ± 15.2 | 70.4 ± 13.0 | **<0.001** (b) |
| Averaged CDR (mean ± Std μm) | 0.42 ± 0.15 | 0.74 ± 0.09 | 0.73 ± 0.09 | **<0.001** (b) |
| VCDR (mean ± Std μm) | 0.40 ± 0.17 | 0.72 ± 0.09 | 0.74 ± 0.10 | **<0.001** (b) |
| sO<sub>2</sub> |  |  |  |  |
| AsO<sub>2</sub> (mean ± Std vol%) | 92.1 ± 7.1 | 88.5 ± 4.7 | 84.5 ± 9.3 | **0.017** (b) |
| VsO<sub>2</sub> (mean ± Std vol%) | 48.4 ± 5.0 | 57.2 ± 8.5 | 56.1 ± 10.8 | **0.009** (b) |
| A-V sO<sub>2</sub> (mean ± Std vol%) | 43.8 ± 9.5 | 31.4 ± 9.3 | 28.3 ± 9.7 | **<0.001** (b) |
| OE (mean ± Std %) | 47.1 ± 7.5 | 35.3 ± 9.5 | 33.6 ± 10.1 | **<0.001** (b) |

(a) Fisher-Freeman-Halton exact test. (b) One-way ANOVA test. (c) t-test. Bold indicates p<0.05

**Figure 2** illustrates the distribution of VIS-OCT measurements, including AsO_2_, VsO_2_, A-V sO_2_, OE, as well as Cirrus measurements of GCL+IPL and cpRNFL in each group. The mean value of AsO_2_ demonstrates a decrease from normal to GS/PPG and further to PG. VsO_2_ significantly increased from normal to GS/PPG and PG, but changed with no significant difference between GS/PPG and PG. A-V sO_2_ and OE exhibit a decreasing trend with severity, indicating reduced oxygen extraction from macular perfusion; these differences were highly significant from normal to GS/PPG (P<0.01) and normal to PG (P<0.001). A-V sO_2_ In Cirrus OCT thickness measurements, GCL+IPL and cpRNFL decrease with severity, consistent with literature. The significant differences occur from normal to GS/PPG and from GS/PPG to PG.

**Figure 2.**
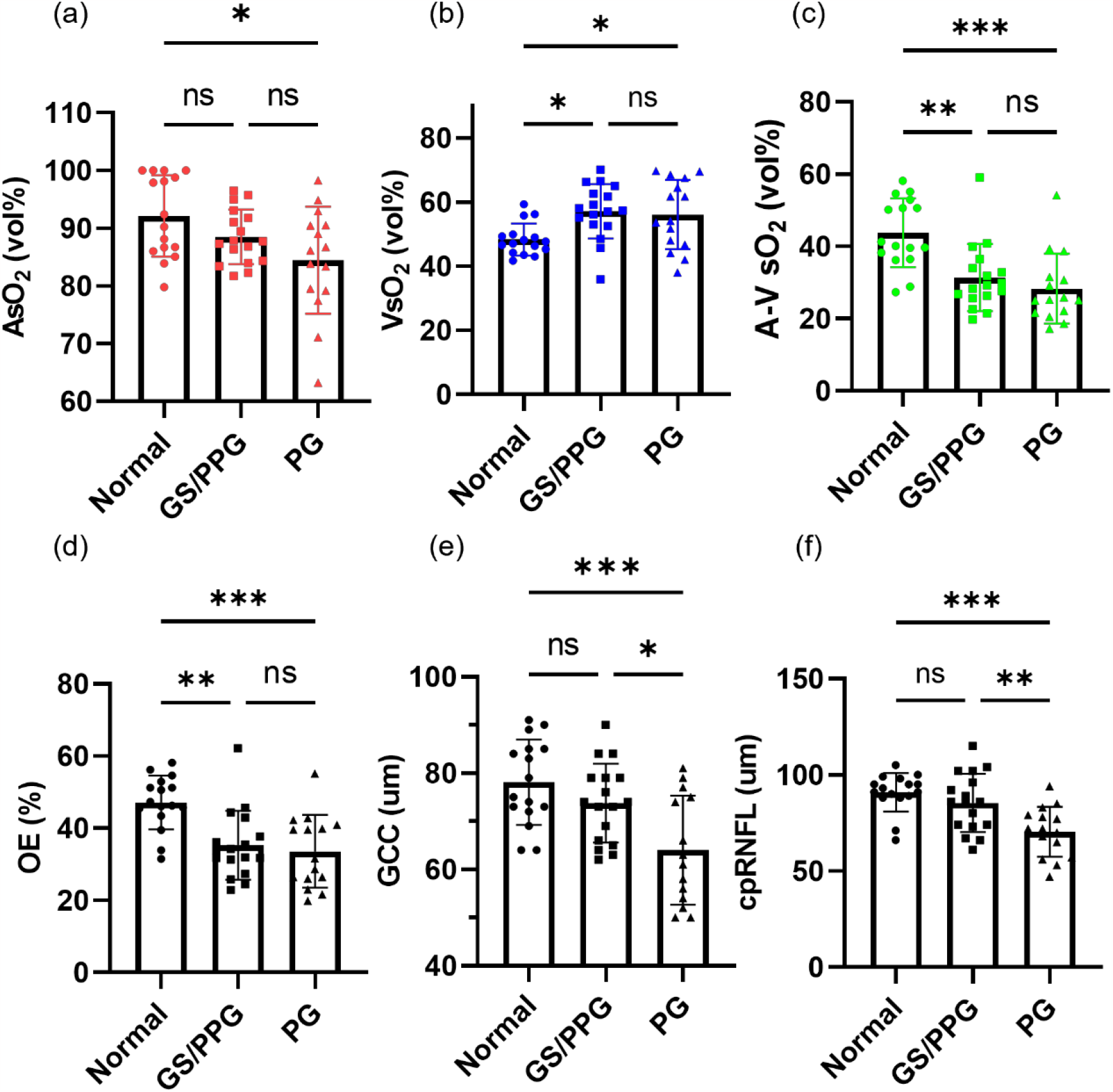
Statistical comparison among normal, GS/PPG and PG groups (a-d) AsO_2_, VsO_2_, A-V sO_2_ and OE among three groups. (e-f) Zeiss OCT thickness including GCL+IPL and cpRNFL. One-way ANOVA was performed first, then multiple comparison within groups was to calculate the p value. ns, not significant, p>0.05. *, p<0.05. **, p<0.01. ***, p<0.001.

### Correlations of key parameters

We performed Spearman correlation among key parameters, including AsO_2_, VsO_2_, A-V sO_2_, OE, GCL+IPL, cpRNFL and MD, as shown in **Table 3**. Within the sO_2_ variables, there is no correlation between AsO_2_ and VsO2_2_. A-V sO_2_ and OE are correlated with AsO_2_ and VsO_2_, as expected. Both GCL+IPL and cpRNFL are significantly correlated with AsO_2_, A-V sO_2_ and OE. MD shows no correlation to the variables mentioned above.

**Table 3.**
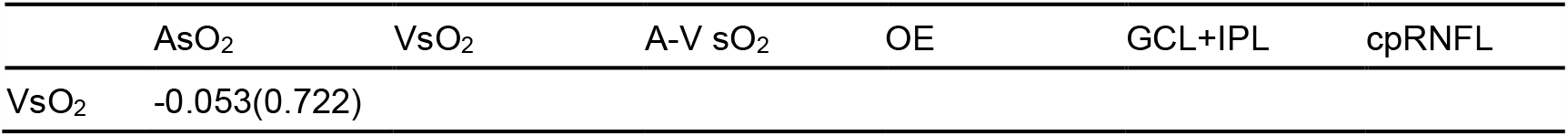
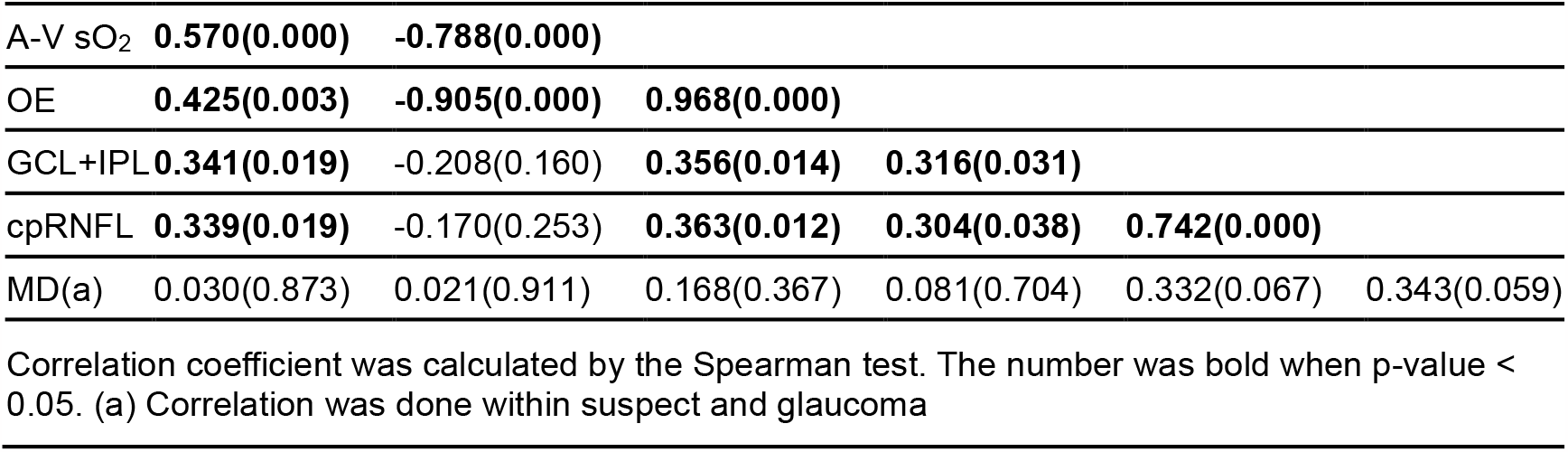
Correlations among groups for key measurements among three groups.

We future investigated the same correlations but only within PG group shown in Table 4. Compared to the result in Table 3, GCL+NFL and cpRNFL are still correlated with AsO_2_ and A-V sO_2_.

**Table 4.** Correlations for key measurements in PG.

|  | As O<sub>2</sub> | VsO<sub>2</sub> | A-V sO<sub>2</sub> | OE | GCL+IPL | cpRNFL |
| --- | --- | --- | --- | --- | --- | --- |
| VsO<sub>2</sub> | 0.354(0.215) |  |  |  |  |  |
| A-V sO<sub>2</sub> | 0.279(0.334) | **-0.714(0.004)** |  |  |  |  |
| OE | 0.130(0.659) | **-0.824(0.000)** | **0.952(0.000)** |  |  |  |
| GCL+IPL | **0.596(0.024)** | -0.077(0.794) | **0.546(0.044)** | 0.460(0.098) |  |  |
| cpRNFL | **0.581(0.029)** | -0.141(0.631) | **0.568(0.034)** | 0.510(0.062) | **0.936(0.000)** |  |
| MD(a) | -0.086(0.771) | -0.222(0.446) | 0.226(0.436) | 0.174(0.553) | 0.407(0.149) | 0.257(0.374) |

Correlation coefficient was calculated by the Spearman test. The number was bold when p-value < 0.05.

In addition, we examined macular sO_2_ by hemifields within PG eyes, as visual damage is preferentially more severe in the lower hemifield than the counterpart (Fig. 3a, 3b), with significantly lower MD. When comparing macular sO_2_ with macular GCL+NFL, a significant correlation is found between AsO_2_ and GCL+NFL in the low hemifield (Fig. 3c, 3d), while the same correlations in the upper hemifield were not significant. This analysis indicates AsO_2_ is correlated with macular RGC loss with confirmed visual field damage.

**Figure 3.**
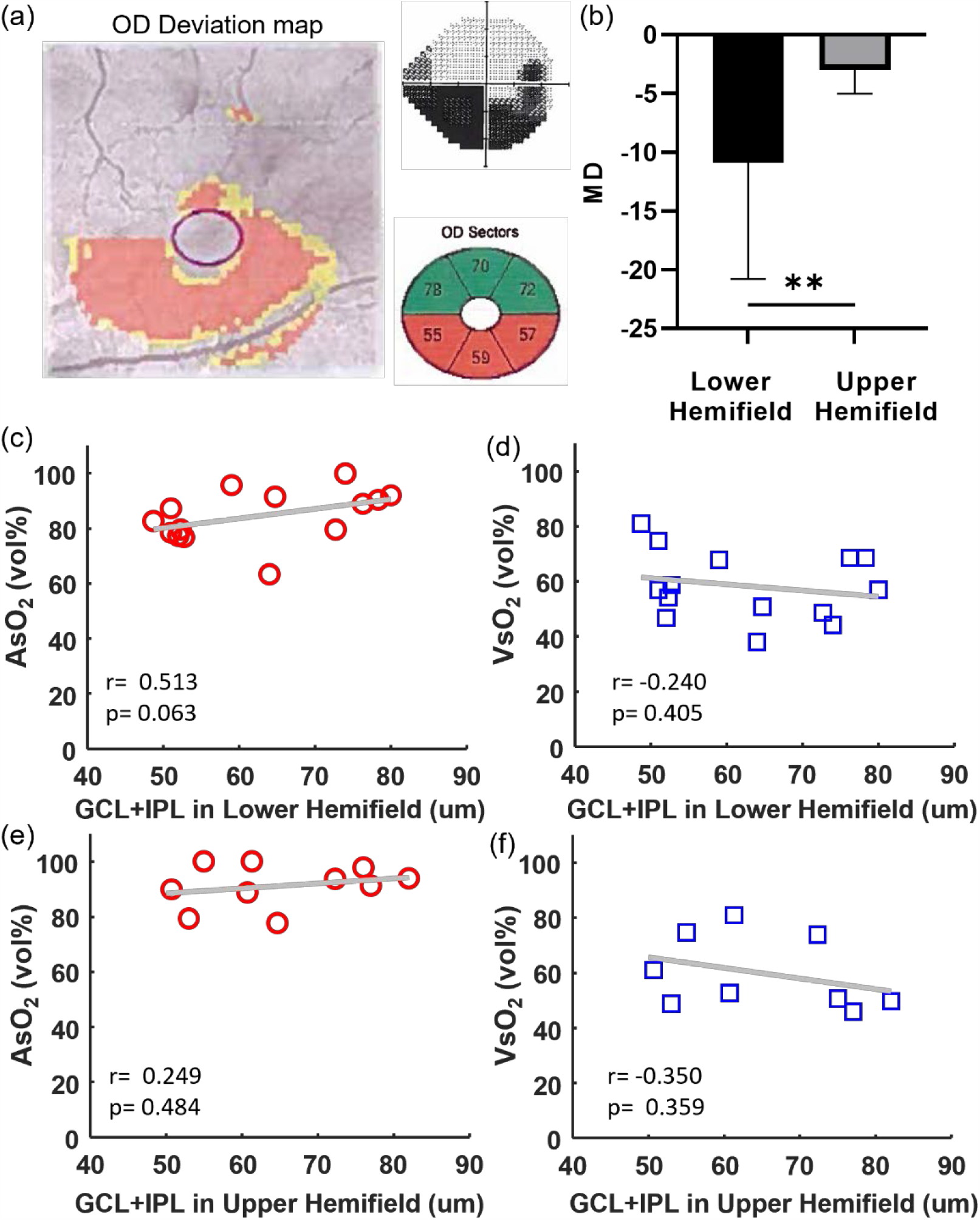
Hemifield analysis in PG subjects. (a) The PG right eye with the deviation map, vision field map and GCL+IPL in different regions and. (b) The MD comparison in the lower and upper hemifields of PG subjects, (c-f) Scatter plot between AsO_2_ and VsO_2_ and GCL+IPL thickness in the lower and upper hemifields in PG subjects. Correlation coefficient and p-value were noted.

### Severity distinguishing

Table 5 illustrates the AUC results of ROC analysis to find the most effective parameter to distinguish among the three groups. All sO_2_ parameters performed better than the GCL+IPL and cpRNFL in distinguishing subjects between normal and GS/PPG, particularly OE. The cpRNFL had the highest AUC area to differentiate GS/PPG from PG.

**Table 5.** Area Under the Curve (AUC) by ROC analysis.

|  | Normal vs GS/PPG | GS/PPG vs. PG | Normal vs. GS/PPG + PG |
| --- | --- | --- | --- |
| AsO<sub>2</sub>, (95% CI) | 0.654 (0.459 to 0.850) | 0.639 (0.439 to 0.839) | 0.692 (0.526 to 0.858) |
| VsO<sub>2</sub>, (95% CI) | 0.824 (0.672 to 0.975) | 0.533 (0.320 to 0.746) | 0.771 (0.639 to 0.904) |
| A-V sO<sub>2</sub>, (95% CI) | 0.827 (0.679 to 0.976) | 0.647 (0.450 to 0.845) | 0.852 (0.741 to 0.962) |
| OE, (95% CI) | **0.849 (0.705 to 0.994)** | 0.557 (0.346 to 0.768) | **0.855 (0.739 to 0.972)** |
| GCL+IPL, (95% CI) | 0.634 (0.442 to 0.826) | 0.737 (0.551 to 0.923) | 0.716 (0.564 to 0.868) |
| cpRNFL, (95% CI) | 0.629 (0.425 to 0.832) | **0.775 (0.613 to 0.936)** | 0.754 (0.608 to 0.900) |
| Receiver operating characteristic (ROC) analysis is based on univariate logistic prediction. The maximum ROC in each column value was bolded. |  |  |  |

## Discussion

This is the first study to investigate macular sO_2_ of glaucoma subjects using the retinal oximetry of dual-channel VIS-OCT. VIS-OCT uniquely allow scanning of the macular arterioles and venules, calculating macular sO_2_, without confounding signals from other layers that plaque the fundus-based oximetry. We found macular sO_2_ is correlated with severity of glaucoma and performs better separating GS/PPG from normal eyes than GCL+NFL and cpRNFL thinning. Within the PG group, AsO_2_ is significantly correlated with GCL+NFL and cpRNFL, which is contributed by the more severely damaged lower hemifield. This correlation analysis strongly indicates that the macular sO_2_ is associated with glaucomatous macular tissue loss. Importantly, macular VsO_2_ and A-V sO_2_ have significant differences when comparing normal with GS/PPG eyes, even prior to detectable thinning and visual field damage.

All existing literatures in the scope of sO_2_ in glaucoma reported global measurements from the major vessels in the parapapillary region around ONH, which is difficult to extrapolate to macular sO_2_ in this presented study^31–38^. Nonetheless, those reports showed rather consistent AsO_2_ in major retinal arterioles, or in rare case higher AsO_2_ in PG subjects. While the inclusion criterion and study population differ, those reports also showed decreased global A-V sO_2_ in glaucoma, underlay by reduced metabolic demand with RGCs loss. Remarkably, we observed a similar reduction of A-V sO_2_, as well as OE at a more localized macular region. Given that RGCs are most abundant in macula, and they are major energy consumer in inner retina, our finding suggests that early RGC/RNFL damage may be manifested with reduction of oxygen metabolism in macular region, correlated with the thinning of GCL+IPL+RNFL. It is worth noting that, in contrary to an unchanged (or slightly increased) global AsO_2_ in previous reports^32–38^, we found that macular AsO_2_ significantly declined with severity among three groups.

Both AsO_2_ and A-V sO_2_ are significantly correlated with GCL+NFL and cpRNFL either among three groups or within PG eyes. In addition, in PG eyes, the correlation is primarily driven by the more severely damaged lower hemifield. To our surprise, we didn’t find significant correlation between VsO_2_ and GCL+NFL or cpRNFL, whether among three groups or within PG eyes. Upon closer examination of the data, VsO_2_ increases significantly from normal to GS/PPG, but remains relatively consistent between two more severer groups. We speculate that VsO_2_ is more significantly impacted in early stage of tissue atrophy in glaucoma, while the AsO_2_ continues to decline along with the progressing severity. A more comprehensive set of including macular blood flow would further elucidate the speculation.

We found no significant correlation of global MD to any macular sO_2_ parameters or GCL+IPL/cpRNFL. We note that 24-2 VFT only has 4 measurements points within 25° center viewing angle in macula, and thus under sampling the macular region which may lead to large variations^16^. Also, as the RGCs are highly redundant in macula, up to 30% of RGCs loss can proceed to visual field damage^10^. Therefore, it is not totally surprising that correlations between MD and other variables are not statistically significant within this study population.

There were several limitations of this study that we would like to address in future studies. First, the cohort size was limited. A larger subject population would be beneficial for multivariable statistical analysis. Second, the implementation of visible light can cause discomfort to some subjects, and it is more susceptible to cataracts and aging eyes. To address this, we will further optimize the imaging device including eye tracking and automatic focusing, which will significantly reduce the exposure to the visible light. Third, the current data processing used manual segmentations for selecting vessel ROIs, as well as assignment of arterioles and venules based on experiences. Automatic algorithms for segmentation and vessel tracing will be developed to streamline the data processing, to achieve real time sO_2_ calculation. Finally, to balance the image depth range, the device used in this study has a relatively short visible light bandwidth of 40nm. Our new-generation VIS-OCT device improved the resolution with wider wavelength bands and expanded the working range^39^. With a more precise extracted spectra, it will help increase the calculation accuracy of sO_2_ in future studies.

In conclusion, we reported the first study of VIS-OCT to investigate the macular sO_2_ in glaucoma subjects. With the high resolution in both axial and lateral directions, VIS-OCT was a powerful tool to characterize macular sO_2_ and identify the significant differences between the normal, GS/PPG and PG subjects. The measurement of sO_2_ can potentially act as a marker to provide the early diagnosis and monitor the progress of glaucoma disease.

## Data Availability

All data produced in the present study are available upon reasonable request to the authors

## Acknowledgments

The study is supported by NIH funding R01NS108464, R01EY032163, and R01EY034607.

## Contribution

JY supervised the project. JW performed the analysis. NS, MF and MD assisted in subject recruitment. All authors have read and approved the final manuscript.

